# Impact of COVID-19 on healthcare workers at a cancer care centre

**DOI:** 10.1101/2021.02.28.21252181

**Authors:** Atika Dogra, Anuj Parkash, Anurag Mehta, Meenu Bhatia

## Abstract

**Background:** The services of front-line health care workers (HCWs) have been paramount in the management of novel coronavirus disease 2019 (COVID-19). Health care professionals have been at high occupational risk of getting disease and even dying of the disease, however; they have been the subject of very limited studies in terms of COVID-19. The objectives of this study are to examine the incidence and the impact of COVID-19 infection among HCWs in terms of recovery, productivity, quality of life (QOL) and post-COVID complications.

**Materials and Methods:** This was a retrospective, questionnaire based study including demographic details, workplace characteristics, symptoms, source/ spread of infection, details of recovery and the consequences of COVID-19 comprising impaired productivity/ QOL, post-COVID-19 complications and others. The data were analyzed by using IBM SPSS software (Version 23, SPSS Inc., Chicago, IL, USA).

**Results and Conclusions:** Out of a total of 1482 employees, 18.3% (271) were laboratory confirmed to have contracted novel contagion during the study period of 5 months. The median age at diagnosis was 29 (range, 21-62) years. Front-line workers and female workers were the most infected personnel with COVID-19. Flu-like symptoms were the most frequently experienced symptoms. The median time for recovery was 20 (range, 2-150) days. The relationship between pre-existing comorbidities and age was highly significant. The QOL and productivity were associated with pre-existing comorbidities, severity of the disease, time for recovery and post-COVID syndrome. More than a half (51.8%) of all HCWs had suffered from post-COVID complications. There was no fatality reported due to COVID-19. The post-COVID complications were related to pre-existing comorbidities, severity of disease, time for recovery and status of recovery. Further research to explore the consequences of COVID-19 is warranted. The general public needs to be aware of symptoms and management of the post-COVID syndrome.

## INTRODUCTION

The novel coronavirus pandemic has become a global health emergency. Worldwide, as of 30 January 2021, 101,561,219 confirmed cases of novel coronavirus disease 2019 (COVID-19) including 2,196,944 deaths, have been reported to WHO from national authorities.^1^ The severe acute respiratory syndrome coronavirus-2 (SARS-CoV-2) is extremely contagious; can cause severe pulmonary infection and respiratory collapse hence early diagnosis of COVID-19 is essential for disease control.^2^ With the rampant increase in the number of infected individuals, the world has seen numerous untimely deaths. Though different fatality ratios have been reported by the countries throughout the world, the highest observed case fatality rate has been reported as 29% from Yemen.^3^

The pandemic has challenged almost every aspect of the health care delivery systems of affected countries putting immense pressure on frontline healthcare workers (HCWs). Every day, they are faced with this daunting reality that they are routinely exposed to the potentially deadly virus. Frontline (HCWs) are facing major difficulties in dealing with patients while keeping themselves and their families safe from COVID-19. The services of front-line HCWs have been paramount in the management of on-going pandemic. The term HCW, defined by WHO includes all people engaged in actions whose primary goal is to improve health.^4^ This includes doctors, nurses, midwives, paramedical personnel, administrative staff, support staff and community employees. With ongoing community transmission and rising disease burden, the health care professionals have been at the high occupational risk of getting disease and at worst, even dying of the disease. A rise in infection and mortality among HCWs may put patients at risk, increases the load on non-infected HCWs and generally hampers the effective delivery of safe and high-quality care. The HCWs have endured various challenges involving long working hours, tiredness, psychological pressure, social stigma and others. Additionally, after having encountered the disease, it is essential for them to resume the duties as early as possible due to the increased workload at working places. The recently emerged data have suggested that many patients have been battling with persisting symptoms even weeks after the onset of initial symptoms of disease which have been named as “Long COVID”^5^ or “post-COVID syndrome”.

The HCWs represent an important but yet understudied population in terms of COVID-19. The objectives of this study are to examine the incidence and the impact of COVID-19 infection among HCWs in terms of recovery, impaired productivity, post-COVID complications and mortality. We have described the patient experience and course of recovery in HCWs having confirmed COVID-19 with a particular emphasis on the post-COVID experience. The understanding of late sequelae of COVID-19 is important in view of very limited information in the public domain.

## MATERIALS AND METHODS

The study was granted a waiver by Institutional Review Board. This was a retrospective, questionnaire based survey and the data of participants had to be utilized in de-identified form so the requirement for informed consent was waived. The first case of COVID-19 infection among the HCWs of our Institute was reported on May 1, 2020. The suspicious cases for COVID-19 were investigated by real-time polymerase chain reaction test and/or cartridge-based nucleic acid amplification test from a combination of naso-oropharyngeal swabs for SARS-CoV-2 as described previously^6^ and/or by Standard Q Covid-19 Ag test following the latest national guidelines of Indian Council of Medical Research and international regulations. The laboratory confirmed COVID-19 positive HCWs during five months (May 1, 2020 to September 30, 2020) were included in the study. A total 271 HCWs were confirmed to have contracted SARS-CoV-2 during above-mentioned period.

The basic details of COVID-19 HCWs were obtained from Human Resource Department of the Institute. The basic information included employees’ IDs, names, e-mail addresses and contact numbers of HCWs. A single page study questionnaire was designed which chiefly included multiple choice questions and a few matrix questions containing textboxes for participants to answer in their own words.

The questionnaire was e-mailed to all the HCWs whose e-mail addresses were available except for 40 HCWs whose e-mail addresses were not obtainable. In the e-mail, the HCWs were first briefed about the study and later were requested to fill their respective questionnaires and submit them back. The questionnaire included demographic characteristics, baseline chronic medical conditions, workplace characteristics (front-line or second-line worker), symptoms present at the time of testing, probable source of infection, further spread of infection, details of recovery and information about the consequences of COVID-19, including impaired productivity/ quality of life (QOL), post-COVID-19 complications and so on. The recovery was defined by the state of becoming free of all symptoms of disease within 3 weeks of symptoms’ onset. The impaired productivity was assessed by the level of efficiency to work and feeling the need for rest which was not the case before. Presently, there is no defined consensus definition for lingering symptoms of COVID-19, we have used the term “post-COVID syndrome”.

The responses of HCWs were awaited for 20 days. The HCWs, whose e-mail IDs were not obtainable, and those who either missed checking or didn’t respond to the e-mail were contacted through telephone. The telephonic survey was started in the first week of November, 2020. The respondents were explained about the study first and their approval for the participation was sought before asking the questions mentioned in the questionnaire. Out of 271, a number of 245 respondents had replied through e-mail and phone. Twenty six people were excluded from the study as they couldn’t be reached out due to non-responsiveness of calls or non-availability of their mobile numbers. The data were collated and anonymized to maintain confidentiality.

### Statistical analysis

The data were analyzed by using IBM SPSS software (Version 23, SPSS Inc., Chicago, IL, USA). The quantitative data were presented by mean (standard deviation [SD]) or median (range) and qualitative data were presented in frequencies/ proportions. The cases with missing records were removed from the analysis. In order to keep the frequencies comparable between the age groups, these were categorized into three groups, 21-30 years, 31-40 years and greater than 40 years. The chi-square test was applied to find out the association between categorical variables. The results were considered statistically significant and highly significant at P <0.05 and P<0.001 respectively.

## RESULTS

Out of a total of 1482 employees, 18.3% (271) had contracted novel contagion during the specified period of 5 months. The mean and median ages at diagnosis were 31.6 (SD, 8.77) and 29 (range, 21-62) years. The most common age group of infected HCWs was between 21-30 years. A majority of the studied cohort to contract COVID-19 was the female population. A major proportion of infected staff (64.5%) belonged to Nursing Department. Table 1 outlines the baseline clinical characteristics of the studied cohort.

**Table 1.** Demographic and clinical characteristics of ovarian cancer.

| **Characteristic (N)** | **Frequency (%)** |
| --- | --- |
| **Age (years) (245)** |  |
| Mean (SD) | 31.6 (8.8) |
| Median (range) | 29 (21-62) |
| **Sex (245)** |  |
| Female | 151 (61.6) |
| Male | 94 (38.4) |
| **Co-morbidities (245)** |  |
| Absent | 222 (90.6) |
| Present | 23 (9.4) |
| **Blood group (237)** |  |
| O+ | 64 (27) |
| O- | 4 (1.7) |
| A+ | 57 (24.1) |
| A- | 2 (0.8) |
| B+ | 83 (35) |
| B- | 4 (1.7) |
| AB+ | 20 (8.4) |
| AB- | 3 (1.3) |
| **Workplace characteristics (245)** |  |
| Front-line workers | 204 (83.3) |
| Second-line workers | 28 (11.4) |
| Others | 13 (5.3) |
| **Symptoms (245)** |  |
| Present | 241 (98.4) |
| Absent | 4 (1.6) |
| **Severity of symptoms (245)** |  |
| Mild | 139 (56.7) |
| Moderate | 68 (27.8) |
| Severe | 34 (13.9) |
| Not applicable | 4 (1.6) |
| **Type of investigation (245)** |  |
| RT-PCR | 239 (97.6) |
| CBNAAT | 2 (0.8) |
| Covid-19 Ag test | 4 (1.6) |
| **Possible source of infection (245)** |  |
| Hospital | 200 (81.6) |
| Community | 24 (9.8) |
| Family | 14 (5.7) |
| Unknown | 7 (2.9) |
| **Further spread (245)** |  |
| No | 193 (78.8) |
| Yes | 52 (21.2) |
| **Hospitalized (245)** |  |
| No | 139 (56.7) |
| Yes | 106 (43.3) |
| **Status of recovery (242)** |  |
| Complete | 124 (51.2) |
| Partial | 118 (48.8) |
| **Impaired quality of life (242)** |  |
| No | 161 (66.5) |
| Yes | 81 (33.5) |
| **Impaired productivity (242)** |  |
| No | 164 (67.8) |
| Yes | 78 (32.2) |
| **Post-COVID syndrome (245)** |  |
| Absent | 118 (48.2) |
| Present | 127 (51.8) |
| **Vital status (245)** |  |
| Alive | 245 (100) |
| Dead | 0 (0) |

Ag, antigen; CBNAAT, cartridge-based nucleic acid amplification test; COVID-19, novel coronavirus disease 2019; RT-PCR, real-time-polymerase chain reaction; SD, standard deviation

### Symptoms

The median duration between onset of first symptom and diagnosis of COVID-19 was 2 (range, 0-22) days. Among all reported symptoms, flu-like symptoms, as defined by National Cancer Institute (National Institute of Health, USA), were most commonly (93.9%) experienced by respondents.^7^ The flu-like symptoms comprise fever, headache, chills, cough, runny nose, sore throat, fatigue, muscle or body aches, nausea, vomiting and diarrhea. Anosmia/ ageusia were experienced by 22.9% HCWs followed by dyspnoea (8.6%), miscellaneous (3.3%) and loss of appetite (2.4%). The miscellaneous symptoms included irritation in eyes, ear pain, skin rash and loss of memory or foggy memory. Four respondents who had tested positive were found to be asymptomatic.

### Source of infection

Two hundred (81.6%) respondents had mentioned that they acquired infection from the workplace. The prime source of infection mentioned were patients (131, 65.5%), followed by coworkers (69, 34.5%). The further transmission of disease from HCWs was reported in 52 (21.2%) cases, which largely spread to family members (42, 80.8%) followed by coworkers (10, 4.1%). Of 245, 106 cases were hospitalized, but all of them did not have severe symptoms and entailed hospitalization. The cause behind this was that most of our hospital’s nurses reside in institutional accommodation and it was made mandatory for them by the administration to get hospitalized in order to quarantine themselves and get treatment and care. The median time of stay at hospital for all admitted personnel was 7 (range, 2-21) days.

### Recovery

The median number of leaves availed by our group of HCWs was 17 days. The median time for recovery was 20 (range, 2-150) days. Ninety three (38.1%) and 72 (29.4%) HCWs had recovered within 17 days and between 18-30 days respectively. Thirty seven (15.1%) respondents took more than 30 days for complete recovery, however; 42 had not recovered completely from the illness at the time of communication with them. The median period of impaired productivity at workplace reported by responders was 30 (range, 5-130) days. The median percentage fall in productivity was reported as 25 (range, 5-80) %. All the patients were alive and there was no single mortality reported due to COVID-19 in our cohort.

### Post-COVID syndrome

One hundred twenty seven (51.8%) HCWs had suffered from post-COVID syndrome. Weakness and fatigue were the predominant symptoms (65.4%) of post-COVID syndrome. The second most commonly reported complication (26.8%) was related to respiratory system followed by neuropsychiatric (14.2%), headache (10.2%), musculoskeletal (6.3%), impaired immunity (4.7%), cardiovascular (4.7%), endocrine system (2.4%) and fever (2.4%). The respiratory symptoms included breathlessness, cough and chest pain. The neuropsychiatric complications comprised of gustatory and olfactory dysfunction, impaired vision and hearing, brain fogging and depression. Musculoskeletal disorders included body ache, joint and knee pain. The impaired immunity symptoms included increased vulnerability to cold and cough. The cardiovascular dysfunction included tachycardia, bradycardia; and 1 patient had developed hypertension as a new manifestation. The post-COVID endocrine dysfunction included the recent development of hypothyroidism, hyperthyroidism, diabetic neuropathy and sudden spurt in glucose levels in diabetics.

### Association between parameters

The symptoms’ severity was significantly associated with all analyzed variables, except for age group, as mentioned in Table 2.The association of symptoms’ severity with recovery shows that the respondents with mild symptoms had largely recovered by the time of survey. The parameter, symptoms’ severity was also significantly related with time for recovery. The majority of respondents with mild symptoms had recovered within 17 days, those with moderate symptoms recovered mostly within 18-30 days; however, those with severe symptoms largely took more than 18 or 30 days. The QOL and productivity were diminished in those individuals whose symptoms were severe, and were least compromised among respondents with mild symptoms.

**Table 2.** Association between severity of symptoms and variables.

| Characteristic (N) | Severity of symptoms |  |  |  | Chi-square | p value |
| --- | --- | --- | --- | --- | --- | --- |
|  | Mild<br>N (%) | Moderate<br>N (%) | Severe<br>N (%) | Not<br>applicable<br>N (%) |  |  |
| **Age group (245)** |  |  |  |  |  |  |
| 21-30 years | 83 (59.7) | 38 (55.9) | 17 (50) | 3 (75) | 9.206 | 0.162 |
| 31-40 years | 32 (23) | 25 (36.8) | 9 (26.5) | 1 (25) |  |  |
| >40 years | 24 (17.3) | 5 (7.3) | 8 (23.5) | 0 (0) |  |  |
| **Status of recovery (243)** |  |  |  |  |  |  |
| Complete | 83 (60.6) | 28 (41.2) | 11 (32.4) | 3 (66.7) | 12.684 | 0.003* |
| Partial | 54 (39.4) | 40 (58.8) | 23 (67.6) | 1 (33.3) |  |  |
| **Time for recovery (243)** |  |  |  |  |  |  |
| ≤17 days | 64 (46) | 19 (27.9) | 6 (17.6) | 3 (75) | 20.452 | 0.007* |
| 18-30 days | 41 (29.5) | 22 (32.4) | 10 (29.4) | 0 (0) |  |  |
| ≥30 days | 14 (10.2) | 13 (19.1) | 10 (29.4) | 0 (0) |  |  |
| Not yet | 19 (13.7) | 14 (20.6) | 8 (23.6) | 1 (25) |  |  |
| **Impaired quality of life (242)** |  |  |  |  |  |  |
| No | 102 (74.5) | 38 (55.9) | 17 (51.5) | 4 (100) | 12.677 | 0.005* |
| Yes | 35 (25.5) | 30 (44.1) | 16 (48.5) | 0 (0) |  |  |
| **Impaired productivity (242)** |  |  |  |  |  |  |
| No | 109 (79.6) | 37 (54.4) | 13 (39.6) | 4 (100) | 28.138 | 0.000** |
| Yes | 28 (20.4) | 31 (45.6) | 20 (60.4) | 0 (0) |  |  |

\* $p < 0.05$ ; \*\* $p < 0.001$

The pre-existing comorbidities were found to be significantly associated with age group and impaired QOL (Table 3). The occurrence of comorbidities was most prevalent in the age group of greater than 40 years and the difference of comobordities’ occurrence between mentioned age group was highly significant. Most of the respondents, who had no pre-existing comorbidity, experienced significantly lower rates of impaired QOL.

**Table 3.** Relationship between co-morbidities and parameters.

| **Characteristic (N)** | **Co-morbidities** |  | Chi-square | p value |
| --- | --- | --- | --- | --- |
|  | **Absent**<br>**N (%)** | **Present**<br>**N (%)** |  |  |
| **Age group (245)**<br>21-30 years<br>31-40 years<br>>40 years | 136 (61.3)<br>60 (27)<br>26 (11.7) | 5 (21.7)<br>7 (30.4)<br>11 (47.9) | 23.744 | .000** |
| **Status of recovery (242)**<br>Complete<br>Partial | 117 (53.2%)<br>103 (46.8%) | 7 (31.8%)<br>15 (68.2%) | 3.653 | 0.056 |
| **Time for recovery (243)**<br>≤17 days<br>18-30 days<br>≥30 days<br>Not yet | 86 (39.1)<br>68 (30.9)<br>30 (13.6)<br>36 (16.4) | 5 (21.7)<br>5 (21.7)<br>7 (30.5)<br>6 (26.1) | 6.918 | 0.068 |
| **Impaired quality of life (242)**<br>No<br>Yes | 151 (68.6%)<br>69 (31.4%) | 10 (45.5%)<br>12 (54.5%) | 4.827 | 0.028* |
| **Impaired productivity (242)**<br>No<br>Yes | 149 (67.7%)<br>71 (32.3%) | 14 (63.6%)<br>8 (36.4%) | 0.152 | 0.696 |

\* $p<0.05$ ; \*\* $p<0.001$

The relationship between time for recovery and all other studied parameters was observed to be statistically significant except for gender (Table 4). It illustrates that the individuals, whose duration for recovery was longer were mainly, from a higher age group and, among those whose QOL and productivity had been affected.

**Table 4.** Relationship between time for recovery and variables.

| Characteristic (N) | Time for recovery |  |  |  | Chi-square | p value |
| --- | --- | --- | --- | --- | --- | --- |
|  | ≤17 days<br>N (%) | 18-30 days<br>N (%) | >30 days<br>N (%) | Not yet<br>N (%) |  |  |
| **Age group (243)** |  |  |  |  |  |  |
| 21-30 years | 63 (69.2) | 43 (58.9) | 13 (35.2) | 21 (50) | 19.728 | 0.003* |
| 31-40 years | 17 (18.7) | 23 (31.5) | 12 (32.4) | 15 (35.7) |  |  |
| >40 years | 11 (12.1) | 7 (9.6) | 12 (32.4) | 6 (14.3) |  |  |
| **Sex (243)** |  |  |  |  |  |  |
| Female | 55 (60.4) | 44 (60.3) | 27 (73) | 24 (57.1) | 2.483 | 0.478 |
| Male | 36 (39.6) | 29 (39.7) | 10 (27) | 18 (42.9) |  |  |
| **Blood group (236)** |  |  |  |  |  |  |
| O+ | 26 (29.9) | 18 (25.4) | 9 (25) | 10 (23.8) | 38.100 | 0.013* |
| O- | 2 (2.3) | 2 (2.8) | 0 (0) | 0 (0) |  |  |
| A+ | 16 (18.4) | 13 (18.3) | 10 (27.7) | 18 (42.9) |  |  |
| A- | 0 (0) | 2 (2.8) | 0 (0) | 0 (0) |  |  |
| B+ | 32 (36.8) | 26 (36.6) | 15 (41.7) | 10 (23.8) |  |  |
| B- | 1 (1.1) | 2 (2.8) | 1 (2.8) | 0 (0) |  |  |
| AB+ | 10 (11.5) | 8 (11.3) | 1 (2.8) | 1 (2.4) |  |  |
| AB- | 0 (0) | 0 (0) | 0 (0) | 3 (7.1) |  |  |
| **Impaired quality of life (240)** |  |  |  |  |  |  |
| No | 81 (90) | 42 (59.2) | 15 (40.5) | 21 (50) | 40.201 | 0.003* |
| Yes | 9 (10) | 29 (40.8) | 22 (59.5) | 21 (50) |  |  |
| **Impaired productivity (242)** |  |  |  |  |  |  |
| No | 85 (94.4) | 42 (59.2) | 16 (43.2) | 18 (42.9) | 53.220 | 0.000** |
| Yes | 5 (5.6) | 29 (40.8) | 21 (56.8) | 24 (57.1) |  |  |

\* $p < 0.05$ ; \*\* $p < 0.001$

The association of post-COVID syndrome with factors was found to be highly significant in many parameters as revealed in Table 5. The occurrence of post-COVID complications or post-COVID syndrome was not statistically different between male and female sub-groups and among different age groups.

**Table 5.** Post-COVID syndrome and parameters.

| Characteristic (N) | Post-COVID syndrome |  | Chi-square | p value |
| --- | --- | --- | --- | --- |
|  | Absent<br>N (%) | Present<br>N (%) |  |  |
| **Age group (245)**<br>21-30 years<br>31-40 years<br>>40 years | 76 (64.4)<br>29 (24.6)<br>13 (11) | 65 (51.2)<br>38 (29.9)<br>24 (18.9) | 5.014 | 0.084 |
| **Sex (245)**<br>Female<br>Male | 69 (58.5)<br>49 (41.5) | 82 (64.6)<br>45 (35.4) | 0.960 | 0.327 |
| **Co-morbidities (245)**<br>Absent<br>Present | 113 (95.8)<br>5 (4.2) | 109 (85.8)<br>18 (14.2) | 7.099 | 0.008* |
| **Severity of symptoms (245)**<br>Mild<br>Moderate<br>Severe<br>Not applicable | 82 (69.5)<br>26 (22)<br>7 (6)<br>3 (2.5) | 57 (44.9)<br>42 (33.1)<br>27 (21.2)<br>1 (0.8) | 20.723 | 0.000** |
| **Time for recovery (243)**<br>≤17 days<br>18-30 days<br>≥30 days<br>Not yet | 89 (76.1)<br>28 (23.9)<br>0 (0)<br>0 (0) | 2 (1.6)<br>45 (35.7)<br>37 (29.4)<br>42 (33.3) | 166.029 | 0.000** |
| **Status of recovery (242)**<br>Complete<br>Partial | 111 (96.5)<br>4 (3.5) | 13 (10.2)<br>114 (89.8) | 179.841 | 0.000** |
| **Impaired quality of life (242)**<br>No<br>Yes | 99 (85.3)<br>17 (14.7) | 62 (49.2)<br>64 (50.8) | 35.422 | 0.000** |
| **Impaired productivity (242)**<br>No<br>Yes | 102 (87.9)<br>14 (12.1) | 61 (48.4)<br>65 (51.6) | 42.897 | 0.000** |

\* $p < 0.05$ ; \*\* $p < 0.001$

## DISCUSSION

To the best of our knowledge, so far, this is the largest report of original research investigating the consequences of SARS-CoV-2 infection among health care professionals in India. The median age of 29 years shows that this cohort has mostly younger population compared to others, who report the higher incidence of COVID-19 in older age groups.^8,9^ It may be due to the reason that the frontline workers at our institute by and large belong to the young age-group. The females were predominantly infected and this is in line with the existing literature.^8^ This may be due to a large number of females workforce as nurses in our Institution. The front-line workers were the ones who were mostly infected which is also related to the preponderance of nurses being front liners. This goes with the fact that the nurses constitute the majority of health professionals and form a significant line of defense.^10^ The pre-existing comorbidities were present in a few patients only, this may be due to the majority of cases being in younger age group.^11^

The highest frequency of flu-like symptoms among reported symptoms may be due to the fact that both, COVID-19 and Influenza are contagious respiratory illnesses and share some common symptoms.^12^ The median duration of 2 days between symptom onset and the diagnosis is shorter than that reported by Mallett *et al*.^13^ This reflects the higher level of awareness among the HCWs compared to general public and the promptness of healthcare system for diagnosis of disease during the pandemic. The number of asymptomatic cases was quite lesser in our study than that reported by others.^14,15^ The predominant occurrence of mild symptoms might be due to the reason that a large number of HCWs were free of pre-existing comorbidities and were in a relatively younger age-group.

Hospital as the main possible source of infection in our patients during pandemic could be due to the maintenance of essential health services and provision of services to all the patients including COVID-19 cases. Only 15.5% of infections were community-acquired which is in contrast with 61.3% reported already.^16^ It may be due to obeying travel restrictions, social distancing rules and quarantine interventions by our staff. Only one-fifth of all cases further transmitted the virus, which again reflects the implementation of strict quarantine policies. The median duration of 17 days as availed leave for sickness was in compliance with the revised guidelines allowing to end home isolation after 17 days of onset of symptoms, if the patient did not have the fever for past 10 days.^17^

Ours may be the first study to report zero COVID-19 deaths. It may be due to cautiousness about the symptoms and disease among employees. Almost half of our HCWs had recovered from COVID-19, however; others still had complaints about the lingering symptoms or the development of new manifestations, even weeks after the symptoms’ onset. Though our COVID-19 survivors had predominantly experienced either mild or moderate illness, the post-COVID syndrome was observed to affect a considerably larger proportion.^18^ The two most common post-COVID symptoms, fatigue and weakness, followed by respiratory system complications were in line with the findings.^19^ The frequencies of post –COVID symptoms have varied with different studied populations.^5,20^ The frequency of brain fogging and cognitive dysfunction (2.4%) in our study was quite lower than reported (85.1%) by Davis et al.^21^ It might suggest the varying outcomes of this virus in different geographical settings.

The association of symptoms’ severity with factor reflects that the individuals with mild symptoms, show higher rates of complete recovery, have recovered early and have lower a propensity of getting affected in terms of QOL and productivity. Our study reveals that the severity of symptoms was not significantly related to increased patient age, the reason of which is not known. This finding is contrary to those reported earlier.^22,23^ Our data also supports the fact that the incidence of comorbidities increases with age.^24^

The median time of recovery from infection in our study corroborated the findings of a Chinese study.^25^ The recovery time is significantly different among different age groups in our study which is similar to the already reported findings.^26^ It reveals that on average, younger individuals are recovering faster from SARS-CoV-2 infection than older patients. The individuals, whose QOL and productivity were not affected, had largely recovered earlier than their counterparts. These characteristics have been studied less as of now, and could barely be found in published literature.

The HCWs with mild symptoms of COVID-19 disease had largely not experienced post-COVID complications. The QOL and productivity were mildly affected in the subset of individuals with no post-COVID syndrome. The subgroup that experienced post-COVID syndrome had predominantly, taken more than 17 days for recovery and partial recovery at the time of survey. Our analysis suggests that post-COVID syndrome includes heterogeneous types of post-infection developments which typically affect multiple organ systems, working and quality of life varying from mild to severe. It is necessary to follow up with the COVID-19 patients to manage ongoing or emerging long-term consequences or complications. Recommended key areas of research into post-COVID syndrome include factors for developing the syndrome, its frequency in different populations, clinically efficient intervention, etc. and lifestyle modification to combat post-COVID complications.

The limitation of our study is the lack of more information about cases that had severe illness and required hospitalization, though the number of such cases was low.

In conclusion, nurses have been affected the most among all healthcare professionals. The QOL and productivity are associated with pre-existing comorbidities, the severity of disease, time for recovery and post-COVID syndrome. Half of all HCWs have been still suffering from post-COVID syndrome and are yet to achieve complete recovery. COVID-19 recovery is linked with the severity of disease. Close attention should be paid to the complications of post-COVID as India records the second highest caseloads with COVID-19 in the world. The general population needs to be aware of post-COVID symptoms and care.

## Data Availability

The data is available in the form of tables

## REFERENCES

1. World Health Organization. COVID-19 Explorer [Internet]. 2021 [Updated 2021 Jan; cited 2021 Jan 30]. Available from https://worldhealthorg.shinyapps.io/covid/

2. Seyed Hosseini E, Riahi Kashani N, Nikzad H, et al. (2020) The novel coronavirus Disease-2019 (COVID-19): Mechanism of action, detection and recent therapeutic strategies. Virology 551: 1–9.

3. Johns Hopkins University & Medicine. Coronavirus Resource Center. Mortality Analyses [Internet]. 2021 [Updated 2021 Feb; cited 2021 Jan 12] Available from https://coronavirus.jhu.edu/data/mortality

4. World Health Organization. Health workers: a global profile, https://www.who.int/whr/2006/06_chap1_en.pdf (2006, xcited 2021 Jan 12).

5. Ladds E, Rushforth A, Wieringa S, et al. (2020) Persistent symptoms after Covid-19: qualitative study of 114 “long Covid” patients and draft quality principles for services. BMC Health Services Research 20(1): 1144.

6. Mehta A, Vasudevan S, Parkash A, et al. (2021) COVID-19 mortality in cancer patients: a report from a tertiary cancer centre in India. PeerJ 9: e10599. DOI 10.7717/peerj.10599

7. National Institute of Health. National Cancer Institute [Internet] 2020 [Updated 2020 Aug; cited on 2021 Jan 17]. Available from https://www.cancer.gov/publications/dictionaries/cancer-terms/def/flu-like-syndrome

8. Bandyopadhyay S, Baticulon RE, Kadhum M, et al. (2020) Infection and mortality of healthcare workers worldwide from COVID-19: a systematic review. BMJ Global Health 5(12): e003097.

9. Kalantari H, Tabrizi A and Foroohi F (2020) Determination of COVID-19 prevalence with regards to age range of patients referring to the hospitals located in western Tehran, Iran. Gene Reports 21: 100910.

10. Al Thobaity A and Alshammari F (2020) Nurses on the frontline against the COVID-19 pandemic: an Integrative review. Dubai Med J 3(3): 1–6.

11. Piccirillo JF, Vlahiotis A, Barrett LB, et al. (2008) The changing prevalence of comorbidity across the age spectrum. Critical Reviews in Oncology/Hematology 67(2): 124–132.

12. Centers for Disease Control and Prevention. Influenza (Flu) [Internet] 2021 [Updated 2021 Jan; cited 2021 Jan 23]. Available from https://www.cdc.gov/flu/symptoms/flu-vs-covid19.htm

13. Mallett S, Allen AJ, Graziadio S, et al. (2020) At what times during infection is SARS-CoV-2 detectable and no longer detectable using RT-PCR-based tests? A systematic review of individual participant data. BMC Medicine 18(1): 346.

14. Al-Qahtani M, AlAli S, AbdulRahman A, et al. (2021) The prevalence of asymptomatic and symptomatic COVID-19 in a cohort of quarantined subjects. Int J Infect Dis 102:285-288.

15. Oran DP and Topol EJ (2020) Prevalence of Asymptomatic SARS-CoV-2 Infection: A Narrative Review. Annals of Internal Medicine 173(5):362–367.

16. Al Maskari Z, Al Blushi A, Khamis F et al. (2021). Characteristics of healthcare workers infected with COVID-19: A cross-sectional observational study. Int J Infect Dis 102: 32– 36.

17. Sharma N. COVID-19 patients can end home isolation after 17 days: Revised guidelines. The Economic Times [newspaper on the Internet]. 2020 May 11 [cited 2021 Jan 23]. Available from https://economictimes.indiatimes.com/industry/healthcare/biotech/healthcare/covid-19-patients-can-end-home-isolation-after-17-days-revised-guidelines/articleshow/75673454.cms

18. World Health Organization. Coronavirus Update 36 [Internet]. 2020 [Updated 2020 Sep; cited 2021 Jan 24]. Available from https://www.who.int/docs/default-source/coronaviruse/risk-comms-updates/update-36-long-term-symptoms.pdf?sfvrsn=5d3789a6_2

19. Centers for Disease Control and Prevention. COVID-19. Long-Term Effects of COVID-[Internet]. 2020 [Updated 2020 Nov; cited 2021 Jan 24]. Available from https://www.cdc.gov/coronavirus/2019-ncov/long-term-effects.html

20. Carfì A, Bernabei R, Landi F, et al. (2020) Persistent symptoms in patients after acute COVID-19. JAMA 324(6): 603–605.

21. Davis H, Assaf G, McCorkell L, et al. (2020) Characterizing Long COVID in an international cohort: 7 months of symptoms and their impact. 10.1101/2020.12.24.20248802

22. Chen T, Dai Z, Mo P, et al. (2020) Clinical Characteristics and Outcomes of Older Patients with Coronavirus Disease 2019 (COVID-19) in Wuhan, China: A Single-Centered, Retrospective Study. J Gerontol A Biol Sci Med Sci 75(9): 1788–1795.

23. Verity R, Okell LC, Dorigatti I, et al. (2020) Estimates of the severity of coronavirus disease 2019: a model-based analysis [published correction appears in Lancet Infect Dis. 2020 Apr 15;:] [published correction appears in Lancet Infect Dis. 2020 May 4;:]. Lancet Infect Dis 20(6): 669–677.

24. Wolff J L, Starfield B, Anderson G (2002) Prevalence, expenditures, and complications of multiple chronic conditions in the elderly. Archives of internal medicine 162(20): 2269–2276.

25. Bi Q, Wu Y, Mei S, et al. (2020) Epidemiology and transmission of COVID-19 in 391 cases and 1286 of their close contacts in Shenzhen, China: a retrospective cohort study. Lancet Infect Dis 20(8): 911–919.

26. Voinsky I, Baristaite G, Gurwitz D (2020) Effects of age and sex on recovery from COVID-19: Analysis of 5769 Israeli patients. The Journal of infection 81(2): e102–e103.

